# Prioritization of therapeutic targets for amyotrophic lateral sclerosis using protein-wide Mendelian randomization analysis

**DOI:** 10.1101/2023.11.22.23298887

**Authors:** Jiawei Geng, Lu-Xi Chen, Chao-Sen Yang, Xixian Ruan, Shixian Hu, Jie Chen, Zhi-Ying Wu

**Affiliations:** Department of Neurology and Research Center of Neurology in Second Affiliated Hospital, and Key Laboratory of Medical Neurobiology of Zhejiang Province, Zhejiang University School of Medicine, Hangzhou, China; Center for Global Health, School of Public Health, Zhejiang University School of Medicine, Hangzhou, Zhejiang, China; Department of Medical Genetics and Center for Rare Diseases, Second Affiliated Hospital, Zhejiang University School of Medicine, Hangzhou, China; Department of Gastroenterology, The Third Xiangya Hospital, Central South University, Changsha, China; Department of Gastroenterology, The First Affiliated Hospital, Sun Yat-Sen University, Guangzhou, Guangdong, China; Institute of Precision Medicine, The First Affiliated Hospital, Sun Yat-Sen University, Guangzhou, Guangdong, China

**Keywords:** amyotrophic lateral sclerosis, Mendelian randomization, drug targets

## Abstract

**Background and Objectives:** Amyotrophic lateral sclerosis (ALS) is a rapidly progressing neurodegenerative disease with an increasing global burden. Available treatments for ALS present marginal efficacy. To identify novel candidate therapeutic targets for ALS, we conducted a proteome-wide Mendelian randomization (MR) study.

**Methods:** We leveraged data from the largest summary statistics for ALS to date (27,205 patients with ALS and 110,881 controls). Genetic instruments of more than 4,000 proteins defined by *cis*-protein quantitative loci (pQTL) genetic instruments on plasma and cerebrospinal fluid (CSF) were obtained from Fenland (discovery, n=10,709), deCODE (replication, n=35,559), and a recently published dataset (replication, n=971). To investigate the causal ALS-associated proteins, proteome-wide Mendelian randomization based on summary-data-based MR (SMR and multi-SNP-based SMR) were performed. Then, two-sample MR analyses using five additional methods were conducted as sensitivity analyses. To further address the linkage disequilibrium bias, heterogeneity in dependent instruments test and colocalization analyses were performed. Steiger filtering and bi-directional MR analyses were conducted to address the potential reverse causality. Four drug target datasets were searched to extract druggability profiles for candidate target proteins. In addition, we carried out a case-control study involving up to 21 patients with ALS and 21 matched controls to assess the protein levels difference in CSF for evidence triangulation.

**Results:** Genetically predicted levels of six circulating proteins were associated with incident ALS in primary SMR analysis. After removing proteins with any linkage disequilibrium bias*, SHBG*, *SIGLEC7, and SIGLEC9* presented consistent associations with ALS risk, supported by medium-to-high colocalization across both plasma pQTL datasets. In CSF, higher level of *SHBG* was also causally associated with the risk of ALS. There was no reverse causality detected. The case-control study using CSF proteomics conducted in our center observed consistent alteration in the levels of *SHBG* and *SIGLEC7* with MR prediction, further suggesting their functionally relevant to ALS as potential druggable targets.

**Discussion:** Combined with the findings from MR and our observational study, we prioritize *SHBG*, *SIGLEC7,* and *SIGLEC9* as drug candidate proteins for ALS, and further studies are needed to verify our findings and elucidate the underlying mechanism.

## Introduction

Amyotrophic lateral sclerosis (ALS) is a rapidly progressing fatal neurodegenerative disease of the central nervous system (CNS). ^1^ Patient with ALS suffer from severe motor dysfunction and respiratory failure, resulting in eventual death within 2-5 years since the initial symptoms onset.^2,3^ ALS is a relatively rare disease, with a global incidence rate of 1.68 per 100,000 person years.^1^ However, with the improvement in diagnosis and worldwide aging, the global burden is increasing.^1, 3^

Despite underlying mechanism of ALS remains largely unknown, available treatments for ALS are suboptimal and primarily relies on symptomatic therapies.^4^ To date, only three disease-modifying agents (Riluzole, Edaravone and AMX0035) established in clinical setting, however, showed marginal efficacy and only applicable to selected population.^1, 5^ Circulating protein plays pivotal roles in biological process, serving as a major source of druggable targets.^6^ Previous studies have identified several ALS-linked proteins.^5^ A recent study revealed pairwise interactions and distinct subgroup aggregation profiles between superoxide dismutase 1 (*SOD1*), TAR DNA-binding protein 43 (*TDP-43*), and ubiquitin-binding protein 62/sequestosome1 (*p62*) in post-mortem ventral spinal cord tissues of ALS patients, which indicated their roles in modulating the formation of proteinopathies and neurodegeration.^7^ A muti-center longitudinal biomarker study involved 108 patients with ALS; and 41 controls with no neurological disease found that cytokines *MCP-1*, *IL-18*, neurofilaments in plasma elevated significantly; And higher level of cerebrospinal fluid (CSF) or plasma neurofilament light chain might associate with fast-progression of ALS.^8^ Therefore, finding more treatments for ALS by identifying circulating proteins with causal evidence might be a promising approach.

Mendelian randomization (MR) used genetic variants as instrumental variables (i.e., protein quantitative trait loci, pQTLs), and has the potential to disclose the causal association between exposure like circulating protein levels and disease outcomes. With the advantages of reducing reverse causation and confounding bias, MR facilitates the understating of the causal mechanisms underlying diseases to further the drug development.^9^ However, the causal association between circulating proteins and ALS is understudied. Therefore, in this study, we leveraged data from large-scale genome-wide association study (GWAS) studies and performed proteome-wide MR and colocalization analyses to identify novel candidate therapeutic targets for ALS.

## Method

### Study design and ethics

The overall study design was shown in **Fig.1**. We first examined the association between protein levels and the risk of ALS using summary-data-based MR test and multi-SNP-based SMR coupled with heterogeneity in dependent instruments. Then, *cis*-MR analyses using five different estimation methods were performed as sensitivity analyses. Followed by colocalization analyses to furtherly account for the potential linkage disequilibrium bias for all identified proteins. Drugabbility of these proteins were appraised based on four datasets. Finally, as external validation, we conducted an investigation on data from our CSF proteomics, including 21 cases and their matched controls, with the purpose to compare the difference in protein levels between groups. All data used in this study were extracted from publicly available summary statistics of GWAS, with ethical permission approved by the corresponding ethics committees and written informed consent obtained from all participants. Detailed data sources and analyses used for the current study are illustrated in **eTable 1**.

**Figure 1:**
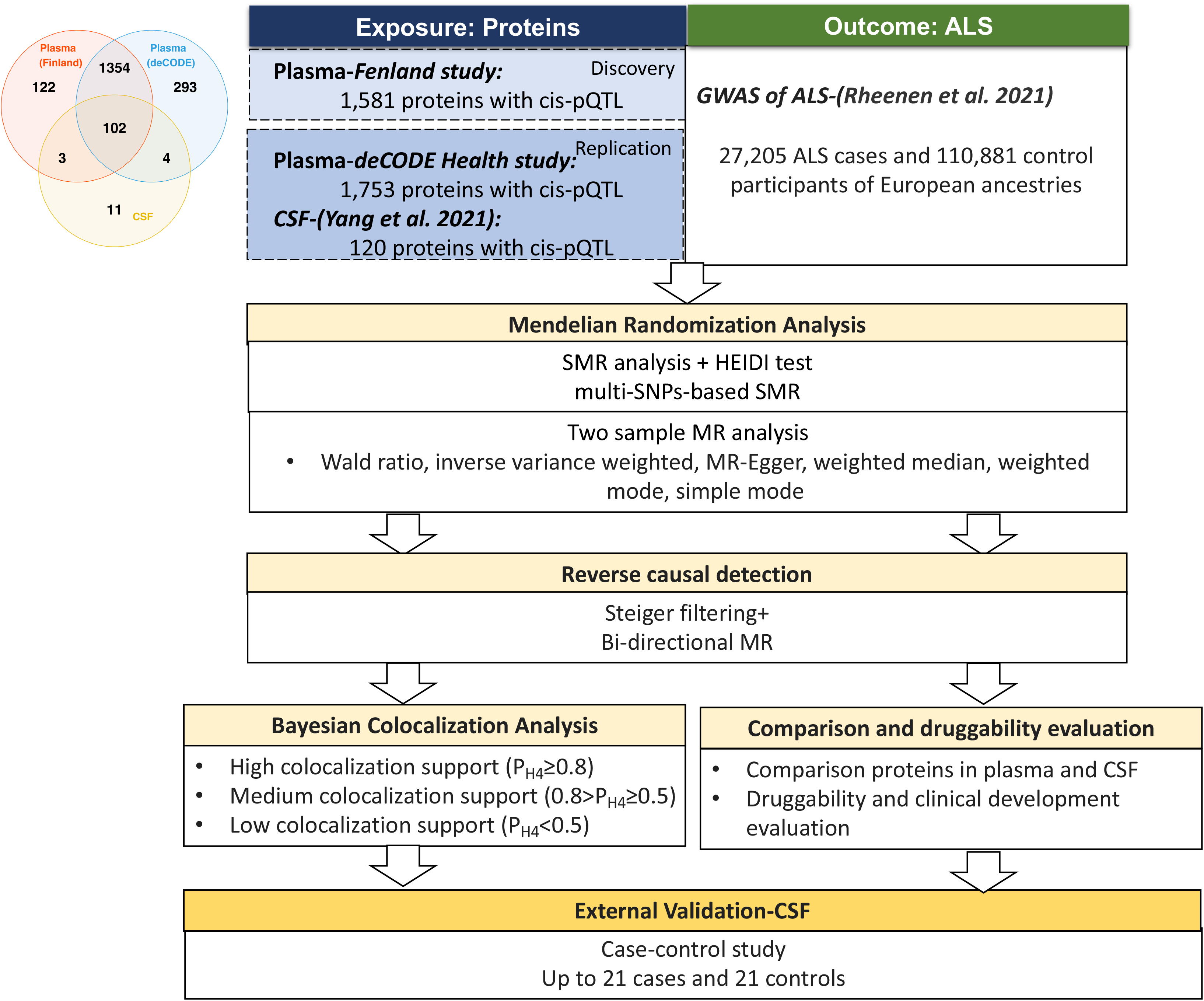
Study design. ALS, Amyotrophic lateral sclerosis; pQTL, protein quantitative trait loci; SMR, summary data-based Mendelian randomization; FDR, False discovery rate; HEIDI, heterogeneity in dependent instruments.

### Protein quantitative trait loci data

#### Plasma

The discovery plasma proteins with quantitative trait loci (*cis*-pQTL) were extracted from the Fenland study, where 4,775 protein targets were measured among 10,708 European-descent individuals. The SomaScan version 4 assay was utilized to measure protein targets, which employed 4,979 aptamers with unique binding affinities. The genetic association was adjusted for age, sex, the first ten principal components and test site. Further details on the data are presented in the original publications.^10^

To validate the effect of identified proteins, we furtherly used another large-scale pQTL dataset conducted among 35,559 Icelanders in the deCODE Health study, with 4,907 aptamers tested and directed towards 4,719 unique proteins (measured with SomaScan version 4).^11^ In deCODE, the genetic association was adjusted for age, sex, and sample age. For these studies, we retrieved SNPs specifically across the levels of the entire proteome (*P*<5X10^-8^).

### Cerebrospinal fluid

Despite CNS drugs delivery is often challenged by the blood-brain barrier and blood-CSF barrier to reach the target disease site^12^, we thus included *cis*-pQTLs from CSF (SomaScan-based, including 971 unique participants) retrieved from the publication of Yang et al.^13^ Due to the limited SNPs obtained meeting genome-wide significance, we, therefore, selected SNPs associated with any protein by a less stringent *P* value threshold (*P*<1X10^-6^) to increase statistical power. MR analysis finally involved 120 proteins with *cis*-pQTLs in CSF.

### ALS GWAS dataset

Summary statistics of ALS obtained from the latest and largest ALS GWAS to date, which contains a total of 138,086 participants of European ancestries (27,205 patients with ALS and 110,881 controls). As described by the original publication,^14^ cases were ascertained according to revised El Escorial Criteria of definite by neurologists. The genetic association was adjusted for principal components (PC)1-PC20.

### Proteome-wide MR

#### Summary data-based Mendelian randomization (SMR) analysis

To identify the genes whose effect on ALS was mediated by the expression of circulating protein, we first conducted the summary data-based Mendelian randomization (SMR) analysis using the top associated *cis*-pQTL.^15^ Linkage disequilibrium (LD) estimation was referenced by 1000 Genomes Project Consortium.^16^ Odd ratios (OR) with corresponding 95% confidence intervals (CI) were estimated to denote per standard deviation (SD) increment in plasma level and 10-fold increment in CSF on the risk of ALS. Given that the blood-CSF barrier might cause the difference in circulating protein levels of plasma and CSF, we also compared the effect sizes of shared pQTLs in these tissues using Spearman correlation analysis. In addition, to further reduce the potential bias of horizontal pleiotropy and also increase the statistical power, the multi-SNPs-based SMR, which extended the SMR method by including multiple SNPs within a 1Mb *cis* region of a probe after removing SNPs exhibited very high LD with the top associated SNP (LD *r*^2^>0.9) was conducted as the sensitivity analysis.^17, 18^ False discovery rate (FDR) multiple testing correction under the Benjamini-Hochberg method was applied, and corrected *P* values less than 0.05 was deemed significant.

To distinguish pleiotropy from the linkage model, we conducted the HEIDI (heterogeneity in dependent instruments) test. *P* values <0.05 were considered as evidence of linkage and we therefore excluded these variants from further analyses. Analyses in this part were performed by the SMR software (Linux version 1.0.3).^15^

#### Two-sample MR

To further assess the causality of association between protein levels and ALS risk, we conducted two-sample *cis*-MR analyses as the sensitivity analyses of SMR. The *cis*-SNP was defined as the SNPs located within 1MB from of the transcription start site of the protein encoding gene.^19^ LD clumping (*r*^2^<0.01) was applied to obtain independent instrumental variable (IV). First, the index *cis*-acting pQTL was retained as the IV for effect estimation. Then, to examine the robustness of estimators, we also conducted MR analysis using all *cis*-acting pQTL that meet the selection criteria for each protein. In two-sample MR, Wald ratio estimation was applied for proteins with single SNP. For those with more than one SNPs, inverse variance weighted (IVW) method was used. Additional MR-Egger^20^, weighted median^21^, weighted mode^22^, simple mode^22^ analyses were performed to indicate the confidence in results where available. We also reported Q statistic to demote the heterogeneity of genetic invariant, and used Egger intercept to detect potential horizontal pleiotropy. We repeated the analyses in Fenland, deCODE, and CSF pQTL datasets. A Benjamini-Hochberg FDR method was conducted, those with adjusted *P* values less than 0.05 were deemed as significant. ‘TwoSampleMR’ package based on R software (version 4.2.0) was used to performed the analyses.

### Reverse causality detection

To account for the potential reverse causality, Steiger filtering was performed to verify the inferenced direction from protein level to incident ALS.^23^ For identified proteins, we also performed bi-directional MR analysis that used ALS as the exposure and proteins levels as the outcomes. Independent SNPs (LD *r*^2^<0.01) associated with ALS clumped at *P*<5 × 10^-8^. Here, we also estimated the casual association used the five two-sample MR analyses described above.

### Colocalization analysis

To further address the potential LD bias underlying MR analysis, Bayesian colocalization analysis using R software (version 4.2.0) with ‘coloc’ package (version 5.2.2) was conducted.^24^ For each locus, 500kb centering on each circulating protein was set. Five exclusive hypotheses to distinguish whether a causal variant is significantly associated with trait 1 (circulating protein) and 2 (ALS), and is shared by these two traits in a region of the genome. Prior probabilities of a variant associated with trait 1, 2, or both 1 and 2 were set at 1X10^-4^, 1X10^-4^, and 1X10^-5^, respectively. Under these assumptions, results of posterior probability for the association of a common variant shared by trait 1 and 2 (H_4_) reached 0.8 was assumed as high-support evidence of colocalization (P_H4_≥0.8, tier 1 target), while those between 0.5 and 0.8 were considered as the medium-support evidence (0.5≥P_H4_>0.8, tier 2 target), and the remaining were regarded as low-support evidence (P_H4_<0.5, tier 3 target).^25^

### Druggability and clinical development evaluation

To assess the druggability and clinical development of each candidate protein, we queried four databases (DrugBank, Dependency Map, the ChEMBL drug discovery database, and the Connectivity Map) supplemented by a druggable gene list^26^ on drug names, clinical development process, and indication conditions. We thus grouped proteins into four categories as follows: 1) approved, 2) in the clinical trial process, 3) preclinical, and 4) druggable.

### External validation

We finally investigated the association between discovered protein levels and ALS in a case-control study. Up to 21 ALS patients and their age, sex, smoking, and alcohol use matched 21 controls were recruited from The Second Affiliated Hospital of Zhejiang University School of Medicine from May 2018 to July 2020. Patients were ascertained with ALS by at least two senior neurologists based on revised El Escorial Criteria. The study obtained ethical permits from the Ethics Committee of the Second Affiliated Hospital in Hangzhou, China, and all participants or their representatives provided written informed consents. The CSF samples (5-8 mL) were collected after discarding the initial 1 mL and were processed and stored in Protein LoBind Tubes (Eppendorf AG, Germany) at a temperature of −80 °C until analysis. Tandem Mass Tag (TMT) proteomics method was used to determine the proteome profiles **(eMethods)**. Student t-test was used to compare the protein levels difference between cases and controls. Two-tailed *P* value <0.05 was deemed as significant.

## Results

### Proteome-wide MR analysis

The overall results in the discovery stage using the data from the Fenland study are displayed in **Fig. 2 and Fig. 3A**. Of the 1,581 unique proteins with available index pQTL signals and ALS outcome, a total of 141 unique proteins reached a nominal significant level (*P_SMR_*<0.05) in the primary analysis **(eTable 2)**. Six protein-ALS pairs survived after FDR correction (*P_FDR_*<0.05) and HEIDI test (*P*≥0.05). Of which, activating transcription factor 6 Beta (*ATF6B*), sex hormone binding globulin (*SHBG*), sialic acid binding lg like lectin 7 (*SIGLEC7*), and sialic acid binding lg like lectin 9 (*SIGLEC9*) showed risk influence on ALS. For per SD increase in genetically predicted plasma protein level, the ORs for ALS were 1.21 (95%CI 1.10-1.33) for *ATF6B*, 1.29 (95%CI 1.13-1.47) for *SHBG*, 1.42 (95%CI 1.19-1.70) for *SIGLEC7*, and 1.06 (95%CI 1.03-1.09) for *SIGLEC9*. Conversely, Tripeptidyl Peptidase 1 (*TPP1*) (OR=0.78, 95%CI 0.69-0.89) and Nucleoside Diphosphate Kinase 4 (*NME4*) (OR=0.84, 95%CI 0.77-0.92) were two proteins identified associated with reduced risk of ALS **(Table 1)**. Secondary analysis from multi-SNPs-based SMR found that the causal association from *ATF6B*, *TPP1,* and *SHBG* with ALS was replicable, and all remaining genes were directionally concordant with estimates generated from primary analysis **(eTable 2)**.

**Figure 2:**
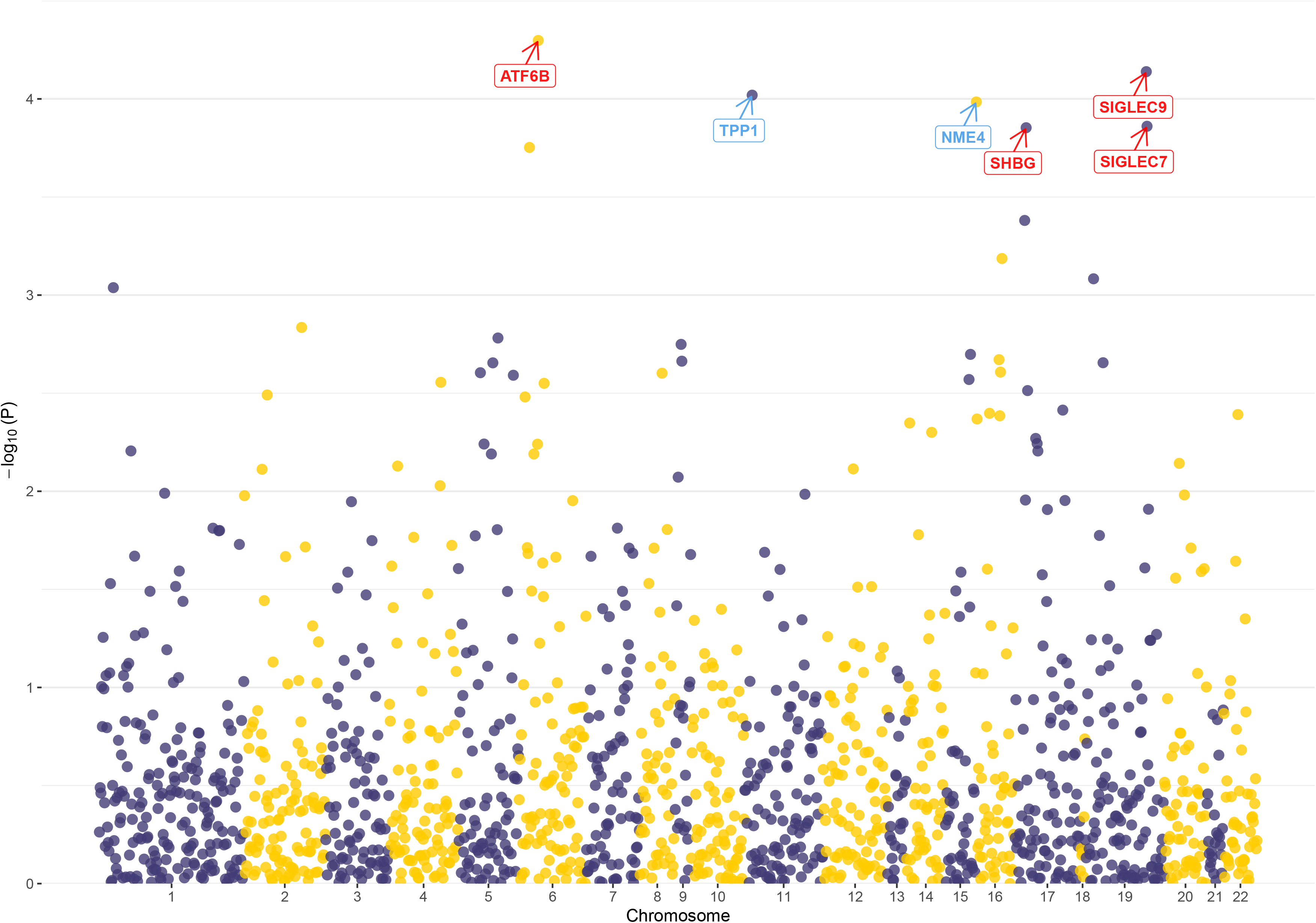
Manhattan plots for associations of genetically predicted circulating protein levels with amyotrophic lateral sclerosis in primary summary data-based Mendelian randomization. Labeled genes are tested with FDR-corrected *P* value <0.05. Genes with positive effects of the circulating proteins on ALS are labeled in red (*ATF6B*, *SHBG*, *SIGLEC7*, and *SIGLEC9*), and genes with negative effects of the circulating proteins on ALS are labeled in blue (*TPP1*, and *NME4*)

**Figure 3:**
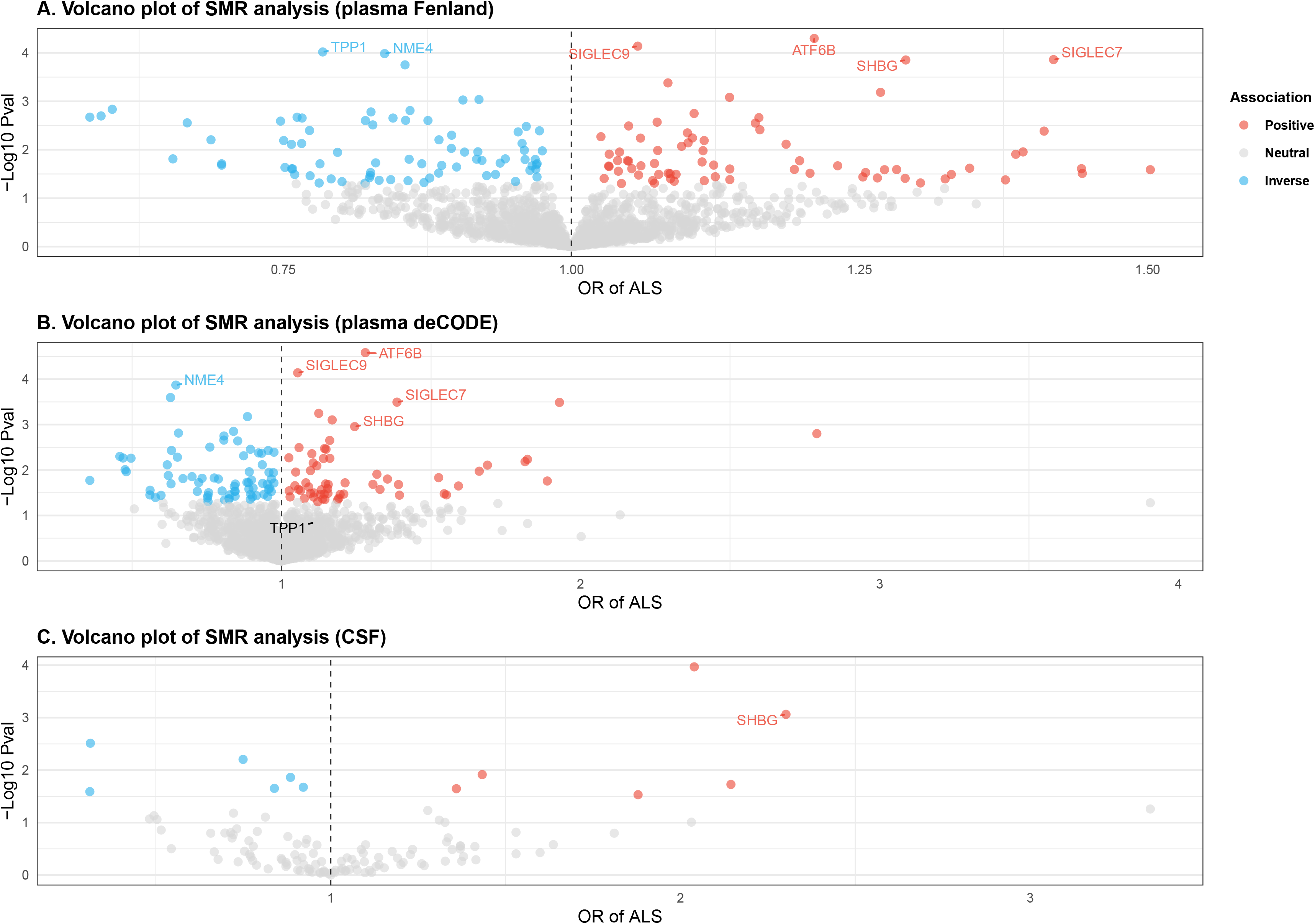
MR results for plasma and CSF proteins and the risk of ALS. Volcano plots of SMR results for (A) 1,581 plasma proteins (Fenland), (B) 1, 753 plasma proteins (deCODE), and 120 CSF proteins on the risk of ALS. Red, blue, and grey points indicate positive (OR >1 and *P*_SMR_>0.05), inverse (OR <1 and *P*_SMR_<0.05), and neutral associations between proteins and ALS. Labeled genes are six genes (*ATF6B*, *TPP1*, *SHBG*, *NME4, SIGLEC7*, *SIGLEC9*) tested with significant FDR-corrected *P* value <0.05 in primary analysis. OR, odd ratios. OR for increased risk of ALS were expressed as per SD increase in plasma protein level and per 10-fold increase in CSF level.

**Table 1:**
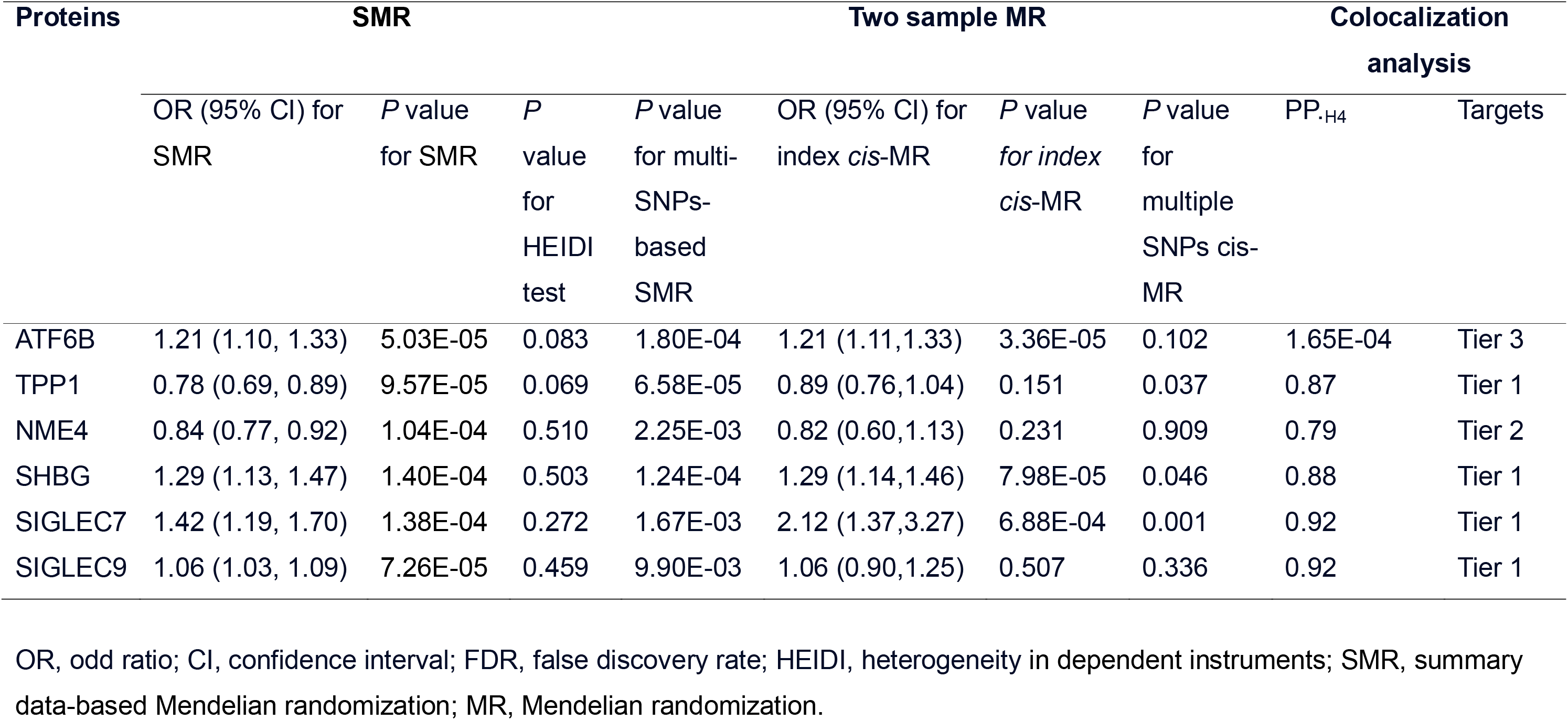
Proteome-wide Mendelian randomization analysis and colocalization of candidate proteins with amyotrophic lateral sclerosis.

Using *cis*-pQTL data in deCODE, five (*ATF6B*, *NME4, SHBG, SIGLEC7,* and *SIGLEC9*) of the six identified proteins appear to be nominally associated with ALS (*P_SMR_<*0.05) in the same direction, suggesting an overall good validity of the findings in Fenland study. *ATF6B* and NME4, however, presented significant HEIDI *P* values, indicating that the associations might attributable to linkage (**eTable 3** and **Fig. 3B**). Notably, the putative causal role of *SHBG*, *SIGLEC7*, and *SIGLEC9* with the risk of ALS observed to be consistent without direct evidence on LD bias. As described in **eTable 3** and **Fig. 3C**, extended analysis in CSF also detected a positive association between *SHBG* expression and ALS. For per 10-fold increase of *SHBG* in CSF, ALS risk would increase 2.3 times (95%CI 1.41-3.76, *P_SMR_*=8.65E-04).

Sensitivity analyses using additional MR analyses furtherly support the findings from SMR. Of note, the causal association of ATF6B and SHBG with the ALS risk passed FDR correction in at least two datasets. Results of index *cis*-MR based on Wald ratio method revealed that the associations between genetically determined higher level of ATF6B and risk of ALS. The odd ratio per SD was 1.21 (95% CI 1.11-1.33) in Fenland (**eTable 4**), and 1.28 (95% CI 1.14, 1.43) in deCODE (**eTable 5**). There was no *cis*-pQTL available for ATF6B in CSF dataset. For SHBG, the OR for ALS per SD increase in genetically predicted levels of proteins were 1.29 (95% CI 1.14-1.46, *P*_FDR_=0.028) in Fenland (**eTable 4**), and in 2.30 (95% CI 1.56-3.39, *P*_FDR_=0.002) CSF dataset (**eTable 6**). SHBG maintained nominally association with ALS risk in deCODE dataset. In Fenland (**eTable 7**) and deCODE datasets (**eTable 8**), where multiple *cis*-pQTL included for IVW estimation, the association between SHBG and ALS risk were directionally consistent with all MR results, with no heterogeneity and pleiotropy discovered (*P* heterogeneity and *P* pleiotropy >0.05). However, in CSF, only one SNPs meet the cis-pQTL selection criteria for SHBG **(eTable 9)**.

### Reverse causality analysis

Results of MR Steiger filtering consistently confirmed the direction of effects between proteins and ALS (**eTable 4, 5 and 6**). Results of reverse MR analyses using the three pQTL datasets presented in Supplementary Table. None of the discovered proteins presented with evidence on the reverse causality (all *P* values >0.05) (**eTable 10**).

### Colocalization analysis

Colocalization analysis involved all circulating proteins that passed FDR correction and HEIDI test in any of the SMR analyses across tissue types **(eTable 11)**. Among these 17 proteins, *NME4*, *SIGLEC7*, and *SIGLEC9* consistently support high evidence of colocalization, showing their potential in sharing the same variant across gene expression with ALS. Of note, *SHBG* provided medium-to-strong evidence for colocalization, and the P_H4_ reached 0.88, 0.52, and 0.97 based on Fenland, deCODE, and CSF studies, respectively. HDGFRP3 (Hepatoma-derived growth factor-related protein 3), a protein significantly associated with ALS in deCODE had medium support of colocalization analysis (P_H4_≥0.5).

### Comparison analysis and druggability evaluation

**Fig 4**. compared MR and colocalization results of potential ALS-associated proteins generated in three databases. Spearman correlation analysis was performed at the protein level to investigate the effect sizes (β) generated from SMR analysis between plasma and CSF pQTL datasets.^27^ We found that the 105 overlapped proteins measured in Fenland and 106 proteins in the deCODE study with CSF were both significantly correlated, with correlation coefficients reached to 0.45 and 0.39 (both *P*<0.001), respectively.

**Figure 4:**
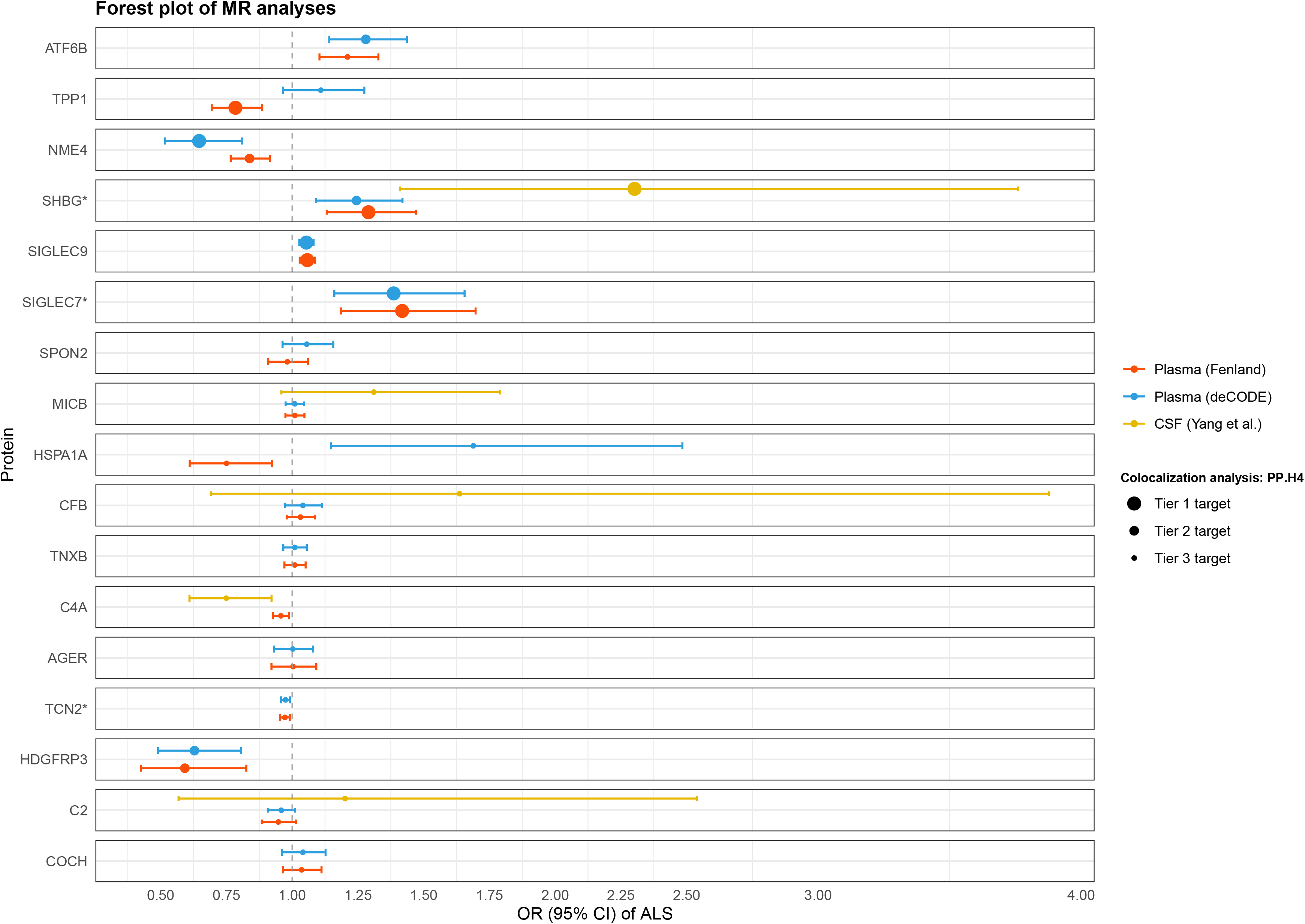
Forest plot of MR analyses of identified proteins in discovery and replication dataset. Results using data from Fenland, deCODE, and CSF are presented with red, blue, and yellow colors, respectively. Proteins found with CSF differences in human subjects are labeled asterisks (SHBG, *SIGLEC*7, and TCN2). OR for increased risk of ALS were expressed as per SD increase in plasma protein level and per 10-fold increase in CSF level. Points size denotes the colocalization probability for H_4_ that is classified by evidence level.

To gain insight into the potentially druggable proteins, we searched the database for all circulating proteins that appeared to be casually associated with ALS in SMR analyses **(eTable 12)**. According to Dependency Map, *ATF6B* and *HDGFRP3* were not currently listed as druggable proteins. *TPP1*, *NME4*, and *SIGLEC9* are druggable but no further information was acquired in searched database. A drug targeting *SIGLEC7* named N-acetyl-alpha-D-glucosamine is currently in the preclinical development phase and was identified in DrugBank. *SHBG* is a major therapeutic target compared to other identified proteins, approved for treating a range of diseases, including cancers, postmenopausal osteoporosis, contraception, immune and allergic disorders, and hormone disorders. However, no drugs pertaining to ALS were developed or in the development based on identified potential causal proteins.

### External validation: a case-control study

To further understand the CSF profiles of candidate proteins, we conducted a sensitivity case-control study comparing the protein levels between patients with ALS and matched controls **(eTable 13)**. The expression of *SHBG* (*P*=0.017), *SIGLEC7* (*P*=0.014), and *TCN2* (*P*=0.020) elevated in ALS-group compared to their matched controls in CSF, suggesting their potential association with the risk of ALS **(eTable 14).**

## Discussion

In the study, we investigated the causal association between more than 4,000 proteins with ALS using 3 proteomic datasets and population-scale genomic data to identify putative therapeutic targets for ALS. In primary MR analysis, six proteins were identified to have potential causal associations with ALS using Fenland pQTL data. Among these, four of these remained directionally consistent between discovery and replication plasma datasets with relatively strong evidence on colocalization analysis. After excluding circulating proteins that exhibited any evidence of linkage disequilibrium bias from HEIDI tests, we finally focused on three proteins, *SHBG*, *SIGLEC7*, and *SIGLEC9*. In particular, *SHBG* and *SIGLEC7* provided supportive evidence in CSF, further suggesting their functionally relevant to ALS and potential druggability. The prioritized actionable target identified in this study is *SHBG,* a circulating glycoprotein mostly produced and secreted by the liver. Its regulation is influenced by hormones and nutritional factors. ^28^ The positive causal association signal of *SHBG* with ALS was replicated in both plasma protein databases by different MR methods, and supplemented colocalization results also support that *SHBG* is likely to share the same variants with ALS. Consistent with our results, genetically determined serum *SHBG* with the risk of ALS was reported by a two-sample MR analysis.^29^ We furtherly added evidence on the causal association between genetically predicted *SHBG* levels in CSF with ALS. In our external validation study, upregulated CSF *SHBG* level was also observed in patients with ALS compared to their matched controls. Although accumulating evidence suggested the increased level of *SHBG* in relation to aging-related neurodegenerative diseases, such as dementias^30, 31^, and multiple sclerosis^32^, observational studies exploring the associated *SHBG* levels in ALS patients were rather limited. One study reported a nonsignificant difference between the group of patients with ALS and their disease controls in serum *SHBG*.^33^ Besides, our result is a contrast to a recent study involved 28 patients with ALS and 22 controls, where serum *SHBG* levels were significantly reduced in ALS group.^34^ The conflicting direction of effects need be interpreted with caution. It is plausible that the selection bias and residual confounding might affect the results since these observational studies are preliminary analyses with a small sample size. In addition, disease progression might also result in discrepant results, which requires further investigation.^35^

Biologic rationale for *SHBG* in elevating the risk of ALS remains to be elucidated. Nevertheless, ALS disproportionally affects males, and it has been hypothesized that testosterone resistance might link with the development of ALS.^29, 33, 36, 37^ *SHBG* is a principal transport protein of testosterone, with roughly 70% of testosterone being bound. ^38^ The neuroprotective function of testosterone in movement and cognition has been widely recognized in vivo and in vitro studies. A study that assessed the concentrations of androgens (testosterone and its main metabolite, dihydrotestosterone) in CSF among 13 patients with ALS and 22 controls observed decreased dihydrotestosterone in the patient group and postulated that malfunctioning transport protein like *SHBG* might hinder testosterone being transferred across the blood-brain barrier.^36^ Experimental studies in Wobbler mice model for ALS have echoed the similar findings. One study revealed a decreased level of testosterone in plasma, indicating its role in motoneuron degeneration.^39^ Another study furtherly confirmed that testosterone treatment slowed ALS progression and improved the rotarod performance of mice. ^40^ Collectively, insights gained above suggested the potential involvement of the endocrine system in etiology as well as the therapeutic value in the treatment of ALS.

Interestingly, two proteins belonging to the *SIGLEC* family (Sialic acid-binding immunoglobulin-type lectins), *SIGLEC7* and *SIGLEC9,* were identified with the concordant direction of effects on ALS with robust evidence on colocalization analysis between both plasma pQTL datasets. *SIGLEC7*, in particular, presented an altered level of CSF in our case-control study. Despite not discovering additional supportive evidence for *SIGLEC9* from either CSF database, a recent BioFINDER study involving 1,591 participants applied different CSF protein panels from Yang et al. implicated shared effects of *SIGLEC9* pQTL (rs2258983) with ALS.^41^ In addition, owing to the close relationship between *SIGLEC9* and *SIGLEC7* (sharing 98% amino acid sequence similarity with the similar expression across immune cells and ligand affinity)^42^, both of these proteins warrant further investigations on their potential in ALS druggability.

Several proteins from the *SIGLEC* family have been linked to neurodegenerative diseases due to their neuroinflammatory properties. ^43, 44^ The role of *SIGLEC7* and *SIGLEC9* in ALS is less explored. *SIGLEC7* and *SIGLEC9* are acknowledged as inhibitory receptors, inducing immune suppression upon binding with ligands. ^45^ These two proteins are rapidly involving CD-33 related *SIGLEC*,^42^ and CD-33 has been implicated in the regulation of microglia activation. ^46^ Emerging evidence links microglial-mediated neuroinflammation to ALS; activated microglia have been observed from neuroimaging and brain tissues from patients with ALS.^47^ It is suggested that modulating the activation of microglial to treat ALS holds promise. ^48^ The underlying mechanism target on *SIGLEC7* and *SIGLEC9* remains to be clarified, and further study and clinical research are needed.

The highlight of the study is a triangulation of findings across MR and an observational case-control study, which strengthened overall confidence in the results. To our knowledge, the study is the first attempt to integrate both pQTL data from plasma and CSF to explore the causal association between circulating proteins and ALS using multiple causal association study methods under the MR framework combined with colocalization analyses. We noted a weak positive correlation in effect sizes generated from MR analyses between plasma and CSF pQTL datasets. This might be explained by the blood-brain barrier and hint at the possible tissue-specific manifestations. Retrieved data from available large-scale GWAS datasets including Fenland, deCODE, and Yang et al.’s study, we were able to perform cross-tissue replication of the findings from the primary analysis. Besides, restricting our analyses among populations of European ancestry might reduce the bias resulting from mixed genetic backgrounds. The study only included *cis*-acting pQTLs, with the direct involvement in the transcription and/or translation of associated genes, it is reduced the bias caused by horizontal pleiotropy. ^49, 50^

The study has some limitations to acknowledge. First, since few *cis*-variants were identified robustly associated with protein levels in CSF, we released the threshold for SNPs selection. Therefore, we should be cautious that the reliability of the effect estimates might be rendered. Second, the external validation involved a limited number of participants, and only controlled for several confounders. However, we recognized quite a few studies examined the casual or observational association of CSF proteins with ALS, further relevant studies are needed to shed more light on ALS pathogenesis. Third, although we used the latest GWAS data from genetic consortia, we did not have access to alternative ALS GWAS to replicate our findings. Further study which includes other and larger datasets with sufficient statistical power is warranted, and involves other ancestries to increase the generalizability. Fourth, despite only *cis*-pQTLs being included, and the HEIDI test and colocalization analysis were conducted to address the LD bias, a potential bias for unknown pleiotropy could not be fully ruled out.

## Conclusions

In summary, consistent positive associations between higher genetically predicted *SHBG* levels and ALS were detected in proteome-wide Mendelian randomization using data from plasma and CSF. By triangulating the finding from both MR and observational studies, we prioritized *SHBG* as the drug candidate protein for ALS, and further studies are needed to elucidate the underlying mechanism. The causal role presented by *SIGLEC7 and SIGLEC9* is also notable, however, further research into the effect of CSF *SIGLEC7 and SIGLEC9* on ALS is warranted.

## Contributions

All authors read and approved the final manuscript.

Jiawei Geng (Conceptualization: Supporting; Methodology: Equal; Formal analysis: Leading; and Writing - original draft: Leading)

Lu-Xi Chen (Conceptualization: Supporting; Methodology: Equal; Formal analysis: supporting; Investigation: Equal; and Writing - original draft: Equal)

Chao-Sen Yang (Investigation: Leading; Methodology: Supporting; and Writing - review & editing: Supporting)

Xixian Ruan (Conceptualization: Supporting; Methodology: Equal; Formal analysis: Supporting; and Writing - review & editing: Supporting)

Shixian Hu (Conceptualization: Supporting; Methodology: Equal; Formal analysis: Supporting; and Writing - review & editing: Equal)

Jie Chen (Conceptualization: Leading; Data curation: Leading; Methodology: Leading; Formal analysis: Supporting; and Writing - review & editing: Equal)

Zhi-Ying Wu (Conceptualization: Supporting; Data curation: Equal; Funding acquisition: Leading; and Writing - review & editing: Equal)

## Supporting information

eTable

## Data availability

Data used in Mendelian randomization study are available from referenced datasets listed in **eTable 1**. Data from the proteomics study are available upon reasonable request.

## Glossary

ALS: Amyotrophic lateral sclerosis
ATF6B: activating transcription factor 6 Beta
CSF: cerebrospinal fluid
CI: confidence interval
CNS: central nervous system
FDR: False discovery rate
HEIDI: heterogeneity in dependent instruments
HDGFRP3: Hepatoma-derived growth factor-related protein
IVW: inverse variance weighted
GWAS: genome-wide association study
LD: Linkage disequilibrium
MR: Mendelian randomization
NME4: Nucleoside Diphosphate Kinase 4
pQTL: protein quantitative loci
SHBG: sex hormone binding globulin
SIGLEC7: sialic acid binding lg like lectin 7
SIGLEC9: sialic acid binding lg like lectin 9
SMR: summary data-based Mendelian randomization
SOD1: superoxide dismutase 1
SD: standard deviation
TMT: Tandem Mass Tag
TDP-43: TAR DNA-binding protein 43
TPP1: Tripeptidyl Peptidase 1

## Acknowledgments

We want to acknowledge investigators and participants in the Fenland and deCODE study, and cited genome-wide association studies for sharing data. We would also thank the participants enrolled in the case-control study for their participation and assistance.

## Supplementary material

### eTables

**eTable 1.** Data sources for studied phenotypes.

**eTable 2.** Associations of plasma proteins with the risk of amyotrophic lateral sclerosis (ALS) in SMR analysis in Fenland study.

**eTable 3.** Associations of plasma proteins with the risk of amyotrophic lateral sclerosis (ALS) in SMR analysis in replicated stage.

**eable 4.** Associations of plasma proteins with the risk of amyotrophic lateral sclerosis (ALS) use index *cis*-pQTL in MR analysis (Fenland).

**eTable 5.** Associations of plasma proteins with the risk of amyotrophic lateral sclerosis (ALS) use index *cis*-pQTL in MR analysis (deCODE).

**eTable 6.** Associations of CSF proteins with the risk of amyotrophic lateral sclerosis (ALS) use index *cis*-pQTL in MR analysis (Yang et al.’s study).

**eTable 7.** Associations of plasma proteins with the risk of amyotrophic lateral sclerosis (ALS) use all *cis*-pQTL in MR analysis (Fenland).

**eTable 8.** Associations of plasma proteins with the risk of amyotrophic lateral sclerosis (ALS) use all *cis*-pQTL in MR analysis (deCODE).

**eTable 9.** Associations of CSF proteins with the risk of amyotrophic lateral sclerosis (ALS) use all *cis*-pQTL in MR analysis (Yang et al.’s study).

**eTable 10.** Reverse association of genetic liability to amyotrophic lateral sclerosis (ALS) with identified proteins.

**eTable 11.** Colocalization analysis on amyotrophic lateral sclerosis (ALS) with relation to identified proteins.

**eTable 12.** Identification of druggable targets.

**eTable 13.** Characteristics of the patients included in the study.

**eTable 14.** Group comparison on identified protein levels in cerebrospinal fluid between amyotrophic lateral sclerosis (ALS) group and matched controls.

**eMethods.**

## References

1. Feldman EL, Goutman SA, Petri S, et al. Amyotrophic lateral sclerosis. Lancet 2022; 400: 1363–1380. DOI: 10.1016/S0140-6736(22)01272-7.

2. Wolfson C, Gauvin DE, Ishola F, et al. Global Prevalence and Incidence of Amyotrophic Lateral Sclerosis: A Systematic Review. Neurology 2023: 10.1212/WNL.0000000000207474. DOI: 10.1212/wnl.0000000000207474.

3. Akçimen F, Lopez ER, Landers JE, et al. Amyotrophic lateral sclerosis: translating genetic discoveries into therapies. Nature Reviews Genetics 2023. DOI: 10.1038/s41576-023-00592-y.

4. Brown RH and Al-Chalabi A. Amyotrophic Lateral Sclerosis. N Engl J Med 2017; 377: 162–172. 2017/07/13. DOI: 10.1056/NEJMra1603471.

5. Mead RJ, Shan N, Reiser HJ, et al. Amyotrophic lateral sclerosis: a neurodegenerative disorder poised for successful therapeutic translation. Nature Reviews Drug Discovery 2023; 22: 185–212. DOI: 10.1038/s41573-022-00612-2.

6. Santos R, Ursu O, Gaulton A, et al. A comprehensive map of molecular drug targets. Nat Rev Drug Discov 2017; 16: 19–34. DOI: 10.1038/nrd.2016.230.

7. Trist BG, Fifita JA, Hogan A, et al. Co-deposition of SOD1, TDP-43 and p62 proteinopathies in ALS: evidence for multifaceted pathways underlying neurodegeneration. Acta Neuropathol Commun 2022; 10: 122. 2022/08/26. DOI: 10.1186/s40478-022-01421-9.

8. Huang F, Zhu Y, Hsiao-Nakamoto J, et al. Longitudinal biomarkers in amyotrophic lateral sclerosis. Ann Clin Transl Neurol 2020; 7: 1103–1116. 2020/06/10. DOI: 10.1002/acn3.51078.

9. Sekula P, Del Greco M F, Pattaro C, et al. Mendelian Randomization as an Approach to Assess Causality Using Observational Data. J Am Soc Nephrol 2016; 27: 3253–3265.

10. Pietzner M, Wheeler E, Carrasco-Zanini J, et al. Mapping the proteo-genomic convergence of human diseases. Science 2021; 374: eabj1541. 2021/10/15. DOI: 10.1126/science.abj1541.

11. Ferkingstad E, Sulem P, Atlason BA, et al. Large-scale integration of the plasma proteome with genetics and disease. Nature Genetics 2021; 53: 1712–1721. DOI: 10.1038/s41588-021-00978-w.

12. Nance E, Pun SH, Saigal R, et al. Drug delivery to the central nervous system. Nature Reviews Materials 2022; 7: 314–331. DOI: 10.1038/s41578-021-00394-w.

13. Yang C, Farias FHG, Ibanez L, et al. Genomic atlas of the proteome from brain, CSF and plasma prioritizes proteins implicated in neurological disorders. Nature Neuroscience 2021; 24: 1302–1312. DOI: 10.1038/s41593-021-00886-6.

14. van Rheenen W, van der Spek RAA, Bakker MK, et al. Common and rare variant association analyses in amyotrophic lateral sclerosis identify 15 risk loci with distinct genetic architectures and neuron-specific biology. Nature Genetics 2021; 53: 1636–1648. DOI: 10.1038/s41588-021-00973-1.

15. Zhu Z, Zhang F, Hu H, et al. Integration of summary data from GWAS and eQTL studies predicts complex trait gene targets. Nat Genet 2016; 48: 481–487. 2016/03/29. DOI: 10.1038/ng.3538.

16. Auton A, Abecasis GR, Altshuler DM, et al. A global reference for human genetic variation. Nature 2015; 526: 68–74. DOI: 10.1038/nature15393.

17. Wu Y, Zeng J, Zhang F, et al. Integrative analysis of omics summary data reveals putative mechanisms underlying complex traits. Nature Communications 2018; 9: 918. DOI: 10.1038/s41467-018-03371-0.

18. Krishnamoorthy S, Li GHY and Cheung CL. Transcriptome-wide summary data-based Mendelian randomization analysis reveals 38 novel genes associated with severe COVID-19. Journal of medical virology 2023; 95: e28162.

19. Sun BB, Maranville JC, Peters JE, et al. Genomic atlas of the human plasma proteome. Nature 2018; 558: 73–79.

20. Burgess S and Thompson SG. Interpreting findings from Mendelian randomization using the MR-Egger method. European journal of epidemiology 2017; 32: 377–389.

21. Bowden J, Davey Smith G, Haycock PC, et al. Consistent estimation in Mendelian randomization with some invalid instruments using a weighted median estimator. Genetic epidemiology 2016; 40: 304–314.

22. Hartwig FP, Davey Smith G and Bowden J. Robust inference in summary data Mendelian randomization via the zero modal pleiotropy assumption. International journal of epidemiology 2017; 46: 1985–1998.

23. Steiger JH. Tests for comparing elements of a correlation matrix. Psychological bulletin 1980; 87: 245.

24. Wallace C, Giambartolomei C and Plagnol V. Package ‘coloc’. 2023.

25. Chen J, Xu F, Ruan X, et al. Therapeutic targets for inflammatory bowel disease: proteome-wide Mendelian randomization and colocalization analyses. EBioMedicine 2023; 89: 104494. 2023/03/02. DOI: 10.1016/j.ebiom.2023.104494.

26. Finan C, Gaulton A, Kruger FA, et al. The druggable genome and support for target identification and validation in drug development. Sci Transl Med 2017; 9 2017/03/31. DOI: 10.1126/scitranslmed.aag1166.

27. Lin J, Zhou J and Xu Y. Potential drug targets for multiple sclerosis identified through Mendelian randomization analysis. Brain 2023; 146: 3364–3372. DOI: 10.1093/brain/awad070.

28. Simó R, Sáez-López C, Barbosa-Desongles A, et al. Novel insights in SHBG regulation and clinical implications. Trends in Endocrinology & Metabolism 2015; 26: 376–383. DOI: 10.1016/j.tem.2015.05.001.

29. Ou YN, Yang L, Wu BS, et al. Causal effects of serum sex hormone binding protein levels on the risk of amyotrophic lateral sclerosis: a mendelian randomization study. Ann Transl Med 2022; 10: 1054. 2022/11/05. DOI: 10.21037/atm-22-1156.

30. Marriott RJ, Murray K, Flicker L, et al. Lower serum testosterone concentrations are associated with a higher incidence of dementia in men: The UK Biobank prospective cohort study. Alzheimer’s & Dementia 2022; 18: 1907–1918. DOI: 10.1002/alz.12529.

31. Muller M, Schupf N, Manly JJ, et al. Sex hormone binding globulin and incident Alzheimer’s disease in elderly men and women. Neurobiol Aging 2010; 31: 1758–1765. 2008/11/22. DOI: 10.1016/j.neurobiolaging.2008.10.001.

32. Safarinejad M. Evaluation of endocrine profile, hypothalamic–pituitary– testis axis and semen quality in multiple sclerosis. Journal of neuroendocrinology 2008; 20: 1368–1375.

33. Militello A, Vitello G, Lunetta C, et al. The serum level of free testosterone is reduced in amyotrophic lateral sclerosis. Journal of the Neurological Sciences 2002; 195: 67–70. DOI: 10.1016/S0022-510X(01)00688-8.

34. Katzeff JS, Bright F, Lo K, et al. Altered serum protein levels in frontotemporal dementia and amyotrophic lateral sclerosis indicate calcium and immunity dysregulation. Scientific Reports 2020; 10: 13741. DOI: 10.1038/s41598-020-70687-7.

35. Campo MD, Pijnenburg YAL, Chen-Plotkin A, et al. Sex Hormone-Binding Globulin (SHBG) in Cerebrospinal Fluid Does Not Discriminate between the Main FTLD Pathological Subtypes but Correlates with Cognitive Decline in FTLD Tauopathies. Biomolecules 2021; 11 2021/10/24. DOI: 10.3390/biom11101484.

36. Sawal N, Kaur J, Kaur K, et al. Dihydrotestosterone in Amyotrophic lateral sclerosis-The missing link? Brain Behav 2020; 10: e01645. 2020/10/14. DOI: 10.1002/brb3.1645.

37. Vegeto E, Villa A, Della Torre S, et al. The Role of Sex and Sex Hormones in Neurodegenerative Diseases. Endocrine Reviews 2020; 41: 273–319. DOI: 10.1210/endrev/bnz005.

38. Li H, Pham T, McWhinney BC, et al. Sex Hormone Binding Globulin Modifies Testosterone Action and Metabolism in Prostate Cancer Cells. Int J Endocrinol 2016; 2016: 6437585. 2016/12/19. DOI: 10.1155/2016/6437585.

39. Gonzalez Deniselle MC, Liere P, Pianos A, et al. Steroid Profiling in Male Wobbler Mouse, a Model of Amyotrophic Lateral Sclerosis. Endocrinology 2016; 157: 4446–4460. DOI: 10.1210/en.2016-1244.

40. Lara A, Esperante I, Meyer M, et al. Neuroprotective effects of testosterone in male wobbler mouse, a model of amyotrophic lateral sclerosis. Molecular Neurobiology 2021; 58: 2088–2106.

41. Hansson O, Kumar A, Janelidze S, et al. The genetic regulation of protein expression in cerebrospinal fluid. EMBO Mol Med 2023; 15: e16359. 2022/12/13. DOI: 10.15252/emmm.202216359.

42. Ibarlucea-Benitez I, Weitzenfeld P, Smith P, et al. Siglecs-7/9 function as inhibitory immune checkpoints in vivo and can be targeted to enhance therapeutic antitumor immunity. Proc Natl Acad Sci U S A 2021; 118 2021/06/23. DOI: 10.1073/pnas.2107424118.

43. Siddiqui SS, Matar R, Merheb M, et al. Siglecs in Brain Function and Neurological Disorders. Cells 2019; 8 2019/09/25. DOI: 10.3390/cells8101125.

44. Siew JJ and Chern Y. Microglial lectins in health and neurological diseases. Frontiers in molecular neuroscience 2018; 11: 158.

45. Jiang KY, Qi LL, Kang FB, et al. The intriguing roles of Siglec family members in the tumor microenvironment. Biomark Res 2022; 10: 22. 2022/04/15. DOI: 10.1186/s40364-022-00369-1.

46. Eskandari-Sedighi G, Jung J and Macauley MS. CD33 isoforms in microglia and Alzheimer’s disease: Friend and foe. Mol Aspects Med 2023; 90: 101111. 2022/08/09. DOI: 10.1016/j.mam.2022.101111.

47. Quek H, Cuní-López C, Stewart R, et al. ALS monocyte-derived microglia-like cells reveal cytoplasmic TDP-43 accumulation, DNA damage, and cell-specific impairment of phagocytosis associated with disease progression. Journal of Neuroinflammation 2022; 19: 58. DOI: 10.1186/s12974-022-02421-1.

48. Clarke BE and Patani R. The microglial component of amyotrophic lateral sclerosis. Brain 2020; 143: 3526–3539. 2021/01/12. DOI: 10.1093/brain/awaa309.

49. Montgomery SB and Dermitzakis ET. From expression QTLs to personalized transcriptomics. Nature Reviews Genetics 2011; 12: 277–282. DOI: 10.1038/nrg2969.

50. Zheng J, Haberland V, Baird D, et al. Phenome-wide Mendelian randomization mapping the influence of the plasma proteome on complex diseases. Nat Genet 2020; 52: 1122–1131. 2020/09/09. DOI: 10.1038/s41588-020-0682-6.

